# Extensive acquisition of carbapenem-resistant *Acinetobacter baumannii* in Intensive Care Unit patients is driven by widespread environmental contamination

**DOI:** 10.1101/2022.05.19.22275186

**Authors:** Emma L. Doughty, Haiyang Liu, Robert A. Moran, Xiaoting Hua, Xiaoliang Ba, Feng Guo, Xiangping Chen, Linghong Zhang, Mark Holmes, Willem van Schaik, Alan McNally, Yunsong Yu

## Abstract

Carbapenem-resistant *Acinetobacter baumannii* (CRAB) is a major public health concern globally. Often studied in the context of hospital outbreaks, little is known about the persistence and evolutionary dynamics of endemic CRAB populations. A three-month prospective observational study was conducted in a 28-bed intensive care unit (ICU) in Hangzhou, China. A total of 3985, 964 and 119 samples were collected from the hospital environment, patients and staff, respectively. CRAB were isolated from 10.75% of collected samples (n = 551) and whole-genome sequenced. The ICU CRAB population was dominated by OXA-23-producing global clone 2 isolates (99.27 % of all isolates) that could be divided into 20 distinct clusters. CRAB was persistently present in the ICU, driven by regular introductions of distinct clusters. The hospital environment was heavily contaminated, with CRAB isolated from bed units on 183/335 (54.63 %) sampling occasions but from patients on only 72/299 (24.08 %) occasions. CRAB was spread to adjacent bed units and rooms and following re-location of patients within the ICU. We also observed that, over the course of this study, three different plasmids had transferred between CRAB strains in the ICU. The epidemiology of CRAB in this setting contrasted with previously described clonal outbreaks in high-income countries, highlighting the importance of environmental CRAB reservoirs in ICU epidemiology. There is an urgent need for targeted infection prevention and control interventions in endemic settings that can address the global threat posed by this against this multidrug-resistant opportunistic pathogen.

## Introduction

Antibiotic-resistant nosocomial infections are a major threat to global public health. *Acinetobacter baumannii* is a Gram-negative coccobacillus that causes severe disease including pneumonia, urinary tract infection, bacteraemia, meningitis, or skin and soft tissue infections, particularly in hospitalised patients.^1^ Carbapenem-resistant *A. baumannii* (CRAB) are found worldwide and are often only sensitive to tigecycline and polymyxins,^2^ severely limiting treatment options. This prompted the World Health Organisation to designate CRAB a priority organism for which novel therapeutics are urgently required.^3^ In the absence of new therapeutic agents, effective CRAB infection prevention and control (IPC) strategies are important to limit the morbidity and mortality it causes in hospitals.^4^ To this end, it is crucial to develop a thorough understanding of the persistence, transmission and evolution of CRAB populations in nosocomial environments.

*A. baumannii* can persist for prolonged periods on hospital surfaces and medical equipment, and colonise patients within 48 hours of admission.^1^ Asymptomatic cutaneous, pharyngeal and gastrointestinal carriage has often been associated with heavy contamination of patients’ immediate environments.^1^ Transmission is thought to be facilitated by hospital staff, shared equipment, airflow and plumbing.^1^ Outbreaks of *A. baumannii* can prove intractable, requiring interventions or changes to infrastructure that impose clinical, logistical, and financial burdens.^5^

Globally, CRAB populations are dominated by two clones, Global Clone 1 (GC1) and Global Clone 2 (GC2), which correspond to ST1 and ST2, respectively, in the Pasteur MLST scheme.^2^ Carbapenem resistance in GC1 and GC2 isolates is most commonly conferred by the *bla*_OXA-23_ carbapenemase gene, located in IS*Aba1*-bounded composite transposons Tn*2006* or Tn*2009* that are usually inserted in the chromosome.^2^ In other cases, *bla*_OXA-23_ or metallo-β-lactamase genes such as *bla*_NDM-1_ are plasmid-borne.^2^ *Acinetobacter* plasmids are genus-specific and have been typed according to the relatedness of their replication initiation proteins.^6^ However, given there are no publicly-available tools or databases for rapidly identifying and typing *Acinetobacter* plasmids, they are often overlooked in genomic studies.

Epidemics of CRAB often occur in high-income countries after a breach of IPC procedures, introducing and spreading a single clone of CRAB within the hospital.^7^ Such clonal outbreaks are typically resolved after outbreak investigation and targeted interventions. Recent studies have demonstrated the utility of whole-genome sequencing (WGS) for high-resolution characterisation of such single-centre outbreaks or populations.^4,8–10^ Endemic hospital *A. baumannii* populations, however, can be composed of multiple phylogenetic clusters.^11,12^ WGS has been employed to assess multi-centre CRAB populations, revealing that individual hospitals harbour multiple clusters and that these may be found across multiple hospitals.^13,14^ These studies have focused on isolates derived from clinical or patient screening specimens. Given its persistence on surfaces, environmental isolates must also be considered in order to understand the distribution and dynamics of CRAB within individual hospitals in endemic countries. A high-resolution assessment of CRAB strain dissemination, cluster evolution, and horizontal gene transfer dynamics requires deep-sampling and WGS of CRAB in a single setting and this provided the motivation for undertaking this study.

Here we describe a prospective observational study of CRAB in an ICU in Hangzhou, China. Over a three-month period, a deep-sampling approach targeted patients, the hospital environment and hospital staff. Isolates were whole-genome sequenced and high-resolution approaches were used to investigate population structure, dynamics of strain movement and dissemination, and horizontal gene transfer events within the ICU.

## Methods

Methods are outlined briefly here. Full details are provided in the supplementary materials.

### Consent and research ethics

Ethical approval and informed consent were obtained by the Sir Run Run Shaw Hospital (SRRSH) Zhejiang University local ethics committee (approval number 20190802-1). This work was part of a study registered as a clinical trial with ClinicalTrials.gov (NCT04310722). The participation of hospital staff was voluntary.

### Study design and sample collection

We conducted a prospective observational study in a 28-bed ICU in SRRSH, Hangzhou, China over a three month period in 2019 (Figure S1). We planned for patients to be sampled at the beginning of the study or on admission to the ward and weekly thereafter, so long as sampling was not perceived to be medically detrimental. Patient samples were routinely collected from oral and rectal swabs, and from nasogastric, nasojejunal, endotracheal, or tracheostomy tube swabs when present. Clinical samples were taken as part of normal medical practice, and all resulting CRAB isolates were collected. Within bed units, equipment and surfaces were swabbed weekly while the surfaces of sinks in patient rooms were sampled fortnightly. Equipment, surfaces, and sinks in communal areas outside patient rooms were sampled monthly. Staff rectal swabs or stool samples were provided in the first week of each month.

### Antibiotic sensitivity testing

Minimum inhibitory concentrations (MICs) for imipenem, meropenem, cefoperazone-sulbactam (1:1 and 2:1), sulbactam, polymyxin B, tigecycline, ciprofloxacin and amikacin were determined using the broth microdilution method in Mueller-Hinton broth (Thermo Fisher Scientific, USA) with 96-well Sensititre plates (TREK Diagnostic Systems Ltd, East Grinstead, West Sussex, UK) at 37°C overnight.

### Sample processing and DNA sequencing

Samples were cultured on Acinetobacter CHROMagar plates containing 2µg/mL meropenem. DNA was extracted from one isolate per culture-positive sample and Illumina sequenced. A subset of 60 isolates were selected based on unique phylogenetic clustering, antibiotic resistance genes and plasmids for long-read sequencing with the Oxford Nanopore GridION.

### Bioinformatic analysis

Sequence reads were trimmed, assembled and assessed for quality. MLST (https://github.com/tseemann/mlst) was used to determine multi-locus sequence types with the Pasteur and Oxford typing schemes.^15,16^ Typing of capsular polysaccharide (KL) and lipooligosaccharide outer core (OCL) synthesis loci was conducted with Kaptive.^17^ AMRFinder was used to identify antimicrobial resistance genes.^18^ A searchable database of all contigs from all Illumina genomes in this collection was constructed and queried with sequences listed in Table S1 using standalone BLAST.^19^

Snippy v4.4.5 (https://github.com/tseemann/snippy) was used to align Illumina reads against a corresponding reference hybrid assembly and generate a core genome alignment. Polymorphic sites were extracted with Gubbins v2.4.0 excluding those that were predicted to occur via recombination.^20^ Phylogenies were constructed from these polymorphic sites using RaxML v8.2.12 with the GTRGAMMA model and autoMRE rapid bootstopping.^21^ The GC2/ST187 population was partitioned into clusters using FastBAPS.^22^ Divergence dating was undertaken with the least-squares method implemented by IQTree v2.0.3, using the previously generated RaxML tree, Gubbins fasta file, and a GTR+G model.^23,24^ SNP-distances were calculated from the Gubbins-filtered polymorphic sites file using SNP-dists 0.6.3 (https://github.com/tseemann/snp-dists).

A nucleotide sequence database, pAci, was created to facilitate *Acinetobacter* plasmid identification and typing. Plasmid types were represented by replication initiation (*rep*) genes or, for types that lack identifiable *rep* genes, by partitioning or transfer-related genes. pAci includes entries for 52 plasmid types or sub-types, and was formatted for use with ABRicate (https://github.com/tseemann/abricate) to enable rapid screening (Table S1).

## Results

### Abundance of CRAB in patients and the patient environment of the ICU

Over the three-month study, a total of 140 patients (102 male; 38 female; median age 70.99 years; interquartile range=63.15 – 85.60) were sampled. The median length of stay from admission to discharge or the end of the study was six days, with an interquartile range of three to 15 days. Samples were taken each Tuesday, except during week 9 due to a national holiday and patients were screened within the first two full days after admission when possible. “Bed units”, each defined as the environmental sites of each bed and its associated equipment (Figurer S1), were sampled on 335/336 (99.70%) planned routine sampling occasions. Patients were sampled on 299/318 (94.03 %) routine occasions when they were present. Fifteen patients stayed in the ICU but were never sampled.

In total, 4949 samples were collected from planned sampling (Table S2). CRAB was isolated from 532 samples (10.75 %): 432/3985 (10.84 %) environmental samples and 100/964 (10.37 %) patient-screening samples. None of the 119 staff samples were CRAB-positive. This amounted to CRAB isolation from complete sampling sets (i.e. the bed unit and the patient in the bed, if present, were both sampled) on 183/316 (57.91 %) occasions. CRAB was isolated on more routine occasions from bed unit environments (183/335, 54.63 %) than patients (72/299, 24.08 %; Fisher’s exact two-tailed P value <0.0001). Bed units yielded more environmental isolates (428/3095, 13.83 %) than communal areas outside bed units (5/80 samples, 6.25 %; Fisher’s exact two-tailed P value 0.0482). Within bed units, samples from ventilators (80/287 samples, 27.87%) and dispensing trolleys (10/39 samples, 25.64%) were most likely to yield CRAB (Table S3). Almost a third (35/125, 28.0%) of patients screened were CRAB-positive during the study, with most of the positive samples originating from oral (32/254, 12.6 %) or rectal (41/289, 14.2%) swabs from 19 and 22 patients, respectively. Additionally, 19 diagnostic clinical isolates were collected from 12 patients.

MICs for six antibiotics were determined for all 551 isolates (Supplementary dataset 1). This confirmed that all isolates were resistant to imipenem and meropenem. All isolates were resistant to ciprofloxacin but sensitive to polymyxin B and tigecycline, and 274 isolates (49.7%) were resistant to amikacin.

### GC2 CRAB dominated the population, characterised by a number of clusters of low diversity isolates

We performed WGS analysis on all 551 isolates and investigated CRAB diversity in the ICU. The majority of isolates, 543 (98.5%), were classified according to the Pasteur MLST scheme as GC2, and four (0.73%) were ST187 (Figure 1), which differed from GC2 by a single nucleotide polymorphism (SNP) in the *rpoB* locus. The remainder were ST46 (n = 2), ST138 (n = 1) and a novel ST (n = 1) that was designated ST1554 after submission to PubMLST. The ST138 isolate has been described previously.^25^ All GC2/ST187 isolates carried the *bla*_OXA-23_ carbapenemase gene in either Tn*2006* or Tn*2009* inserted in one of five chromosomal positions (Figure 1). Other acquired antibiotic resistance genes found in GC2/ST187 isolates conferred resistance to sulphonamides, aminoglycosides, tetracycline or macrolides and were in variations of the AbGRI1, AbGRI2 and AbGRI3 chromosomal resistance islands (Figure 1). IS*26*-mediated deletion events have been responsible for the loss of resistance genes from these islands (Figure 1). Amikacin resistance was strongly associated with the presence of the *armA* aminoglycoside resistance methylase gene in AbGRI3. All 271 *armA*-containing isolates were amikacin resistant, but the remaining three amikacin-resistant isolates did not contain *armA*.

**Figure 1.**
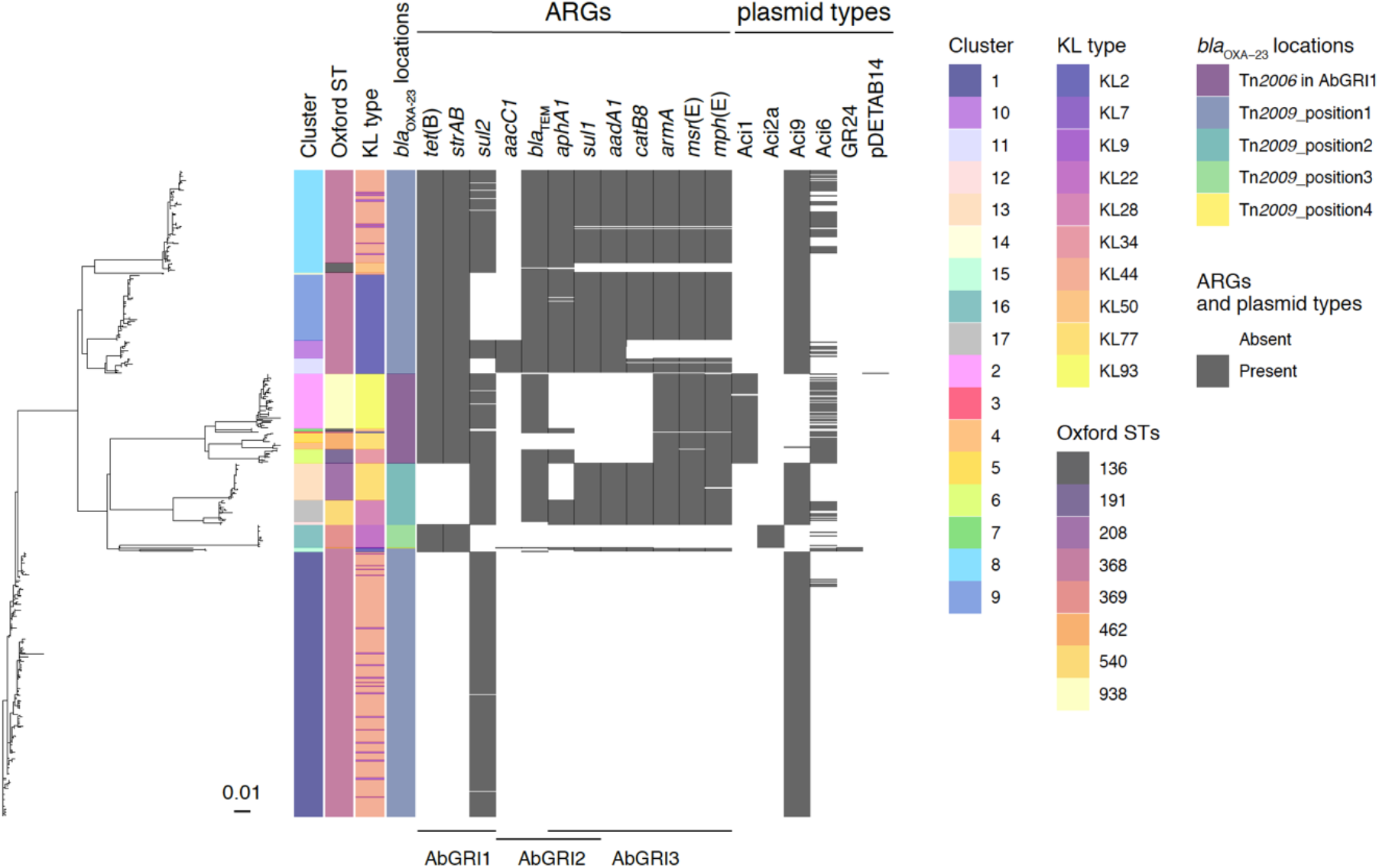
Characterisation and comparison of GC2/ST187 CRAB found in the ICU. Evolutionary relationships of the 547 GC2/ST187 isolates from the ICU; coloured tracks adjacent to the phylogenetic tree tip show the cluster designation, Oxford MLST types, KL types and different positions of *bla*_*OXA-23*_ genes; presence of each antibiotic resistance gene and plasmid type is shown in grey and their absence is in white.

Diversity of the GC2/ST187 CRAB population was explored in terms of Oxford MLST and typing of the capsular polysaccharide (KL) and lipooligosaccharide outer core (OCL) synthesis loci. Amongst the 547 isolates, eight Oxford STs and ten KL were identified (Figure 1). All isolates were OCL1. In 489 isolates, there were two copies of the *gdhB* gene used in the Oxford MLST scheme, a phenomenon which has been noted previously.^26^ In all instances, one copy was identified as allele 189 and the other was a variable allele used to identify the multilocus sequence type. K loci were assigned with variable match confidence levels and assignment problems were reported for some isolates. In many cases Oxford ST and/or KL profiles were incongruent with the phylogenetic population structure (Figure 1). Both KL7 and KL28 occurred within the same phylogenetic clade while ST_OX_136, ST_OX_208, ST_OX_540, KL2, KL7, KL9, KL28 and KL77 were assigned to isolates from multiple polyphyletic clades. The incongruence of Oxford STs and KL types with the GC2 phylogeny can be explained by the presence of Oxford MLST genes *gyrB* and *gpi* (which is also part of the K locus) in genomic regions that had been subject to recombination, with extensive recombination occurring in all strains sequenced in the capsule locus (Figure 2). Neither Oxford MLST nor K/OCL typing were able to adequately distinguish subpopulations of GC2/ST187 circulating in the ICU for epidemiological typing purposes.

**Figure 2:**
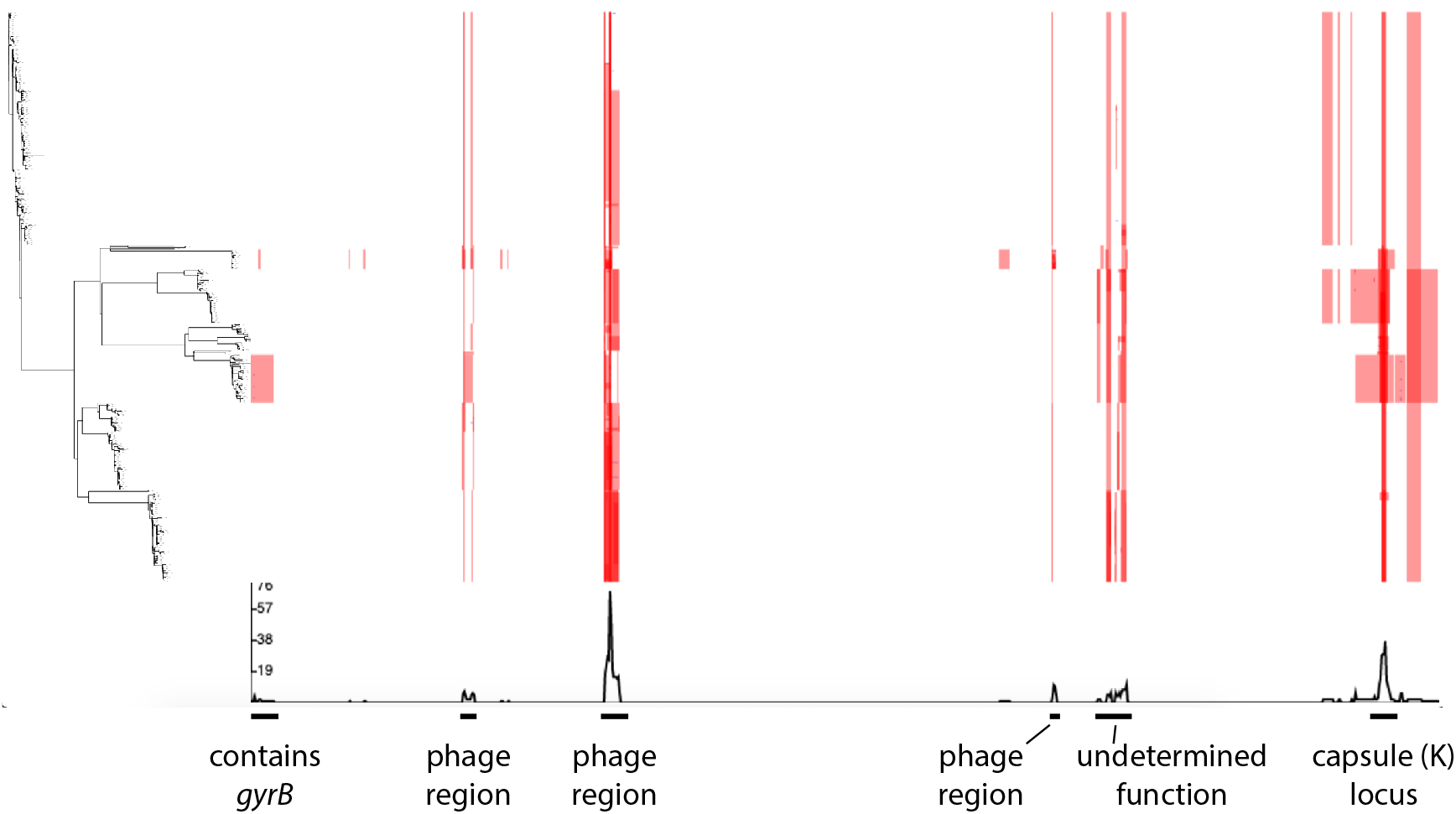
Recombination occurring in the sequenced GC2/ST187 CRAB isolate set. The plot shows recombination as detected using Gubbins on all CRAB genome sequenced in the study, superimposed on the data set phylogeny. The red shading indicates recombining regions of the genome, annotated underneath with functional gene annotation.

Phylogenetic and population genetic analysis of the GC2/ST187 isolates revealed that this population had a maximum of 194 core-genome SNPs between isolates. These were used to partition the population into 17 distinct GC2/ST187 clusters (each prefixed with “C”) using FastBAPS (Figure 1). Published mutation rates for GC2 *A. baumannii* estimate a rate of approximately 24 substitutions per year for within host evolution and approximately 10 substitutions per year for GC2 spreading between hosts.^10,27^ The minimum SNP distance between our 17 clusters ranged from 12 to 117 SNPs indicating that the diversity seen across our GC2 isolates has not accumulated within the ICU from a single source introduction during the sampling period (Table S3). Dating analysis using IQtree supported this assertion, (Table S4) with the most recent common ancestors (MRCA) between clusters predicted to occur prior to the start of this study (from 31/03/1992 to 19/04/2016). Combining these data with published mutation rates and IQTree dating shows that these clusters did not arise from one another over the course of this study. Rather, each of these clusters was introduced separately or circulating independently within the ICU setting during our sampling period. The median within cluster SNP distance across the 17 clusters ranged from 0-8 SNPs with the exception of cluster 15 which has a within cluster median SNP distance of 76 SNPs (Table S3).

Of 53 patients screened within two full days of ICU admission, four yielded CRAB from oral, rectal or indwelling tube swabs. Three of these samples represented the first or only appearance of ST138, cluster 3 and cluster 9, strongly suggesting that they were introduced to the ICU with their respective patients. The fourth patient also yielded cluster 9, which had not been detected in the ICU for six weeks prior to their admission. This suggested that cluster 9 was introduced to the ICU on two occasions by two different patients. Additionally, three patients yielded CRAB from sputum samples collected for clinical purposes within 48 hours of ICU admission. Two of these represented the first appearances of clusters 14 and 16. Thus, at least five CRAB clusters appear to have been introduced to the ICU with patients (Table S5).

### Extensive strain spread in the ICU and acquisition by patients was driven by environmental contamination

Six CRAB clusters (C3, C12, C14, and ST46, ST138 and ST1554), each with a maximum of three isolates and associated with a single patient were considered sporadic. The remaining 14 clusters were found in between two and 49 different patients or their bed unit environments (Table S6). For these clusters, the median number of cgSNPs was between 0-8 SNPs, and the distribution of small cgSNP frequencies around a mode (Figure S2) was clearly indicative of their recent and ongoing spread within the ICU.

Cluster 1, represented by 224 isolates, was isolated in each week of the study period, and at least once from each of the patient rooms in the ICU. This dominant and seemingly endemic cluster provided an opportunity to examine the dynamics of a clonal population that persisted in the ICU environment for the entirety of the study period. A maximum-likelihood core-genome phylogeny revealed that all but one isolate in cluster 1 differed by ≤7 SNPs from the most basal isolate (Figure 3A). The distribution of cluster 1 throughout the study indicates it persisted and spread within the ICU (Figure 3B) and was introduced on multiple occasions from a broader nosocomial population. For the first seven weeks, the cluster 1 population was dominated by isolates that differed by ≤3 SNPs from the basal isolate (orange shades in Figure 3), while isolates that differed by 4-7 SNPs (pink/purple shades in Figure 3) appeared sporadically. In the final six weeks of the study, isolates 4-7 SNPs from basal dominated the population, centred around the environment of room 5 (Figure 3B). Most cluster 1 isolates in room 5 at this time were associated with patient 91, who yielded cluster 1 from a clinical sputum sample within 48 hours of admission to the ICU. Cluster 1 was isolated from further patient 91 sputum and oral/rectal swabs in subsequent weeks, as well as from the room 5 environment. The most distantly related cluster 1 isolate, which differed from the basal isolate by 19 SNPs (Figure 3A), was isolated from a stethoscope in patient 91’s bed unit environment in this period, as were isolates that differed from the basal by ≤3 SNPs. In this and other instances, simultaneous isolation of different sub-populations from the same bed units suggest that cluster 1 had persisted and diversified in this hospital for an extended period of time and was introduced into the ICU on multiple occasions.

**Figure 3.**
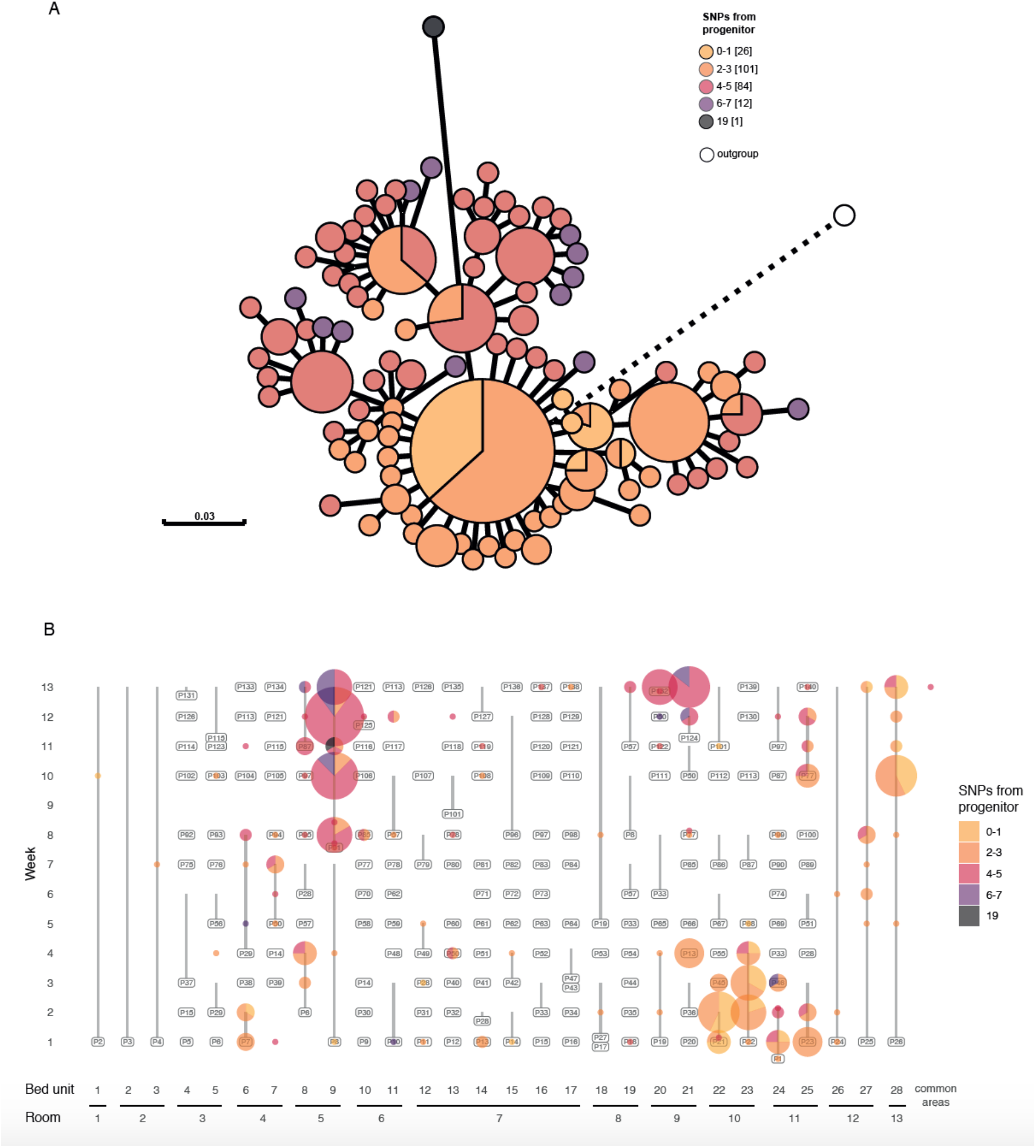
Introduction and spread of cluster 1 within the ICU as measured by whole genome SNPs. A) Maximum-likelihood core-genome phylogeny of all 224 cluster 1 isolates using a cluster 11 isolate (DETAB-E47) as an outgroup. Visualised in GrapeTree and rooted on the outgroup, shown in white. Colours show the number of SNPs from the most basal cluster 1 isolate, with up to 7 cgSNPs in the nosocomial population, and 19 cgSNPs in the most distal isolate (DETAB-E290 which may have been introduced into the ICU separately as it was found in a single patient on their admission to the unit). B) Spatiotemporal distribution of cluster 1 isolates, coloured as in panel A.

In contrast to cluster 1, cluster 2 was introduced to the ICU during the study period, facilitating an examination of its subsequent dissemination dynamics (Figure 4). The first appearance of cluster 2 was in a rectal swab from patient 97 in week 8, four days after they had been admitted to bed unit 16. The absence of cluster 2 prior to their admission strongly suggests that it was introduced to the ward with patient 97 (Table S5). On the same day that cluster 2 was first isolated, patient 96 was admitted to bed unit 15 and shown to be CRAB-negative via oral and rectal swabs. By the next sampling date, patient 97 had been moved to bed unit 8 in room 5, but cluster 2 had persisted in room 7, where it was found in the environments of bed units 12, 14, 15 and 16. Cluster 2 was also isolated from patient 96 oral and rectal swabs at this time, indicating that they had acquired CRAB within two weeks of admission to the ICU. Patient 96 continued to harbour cluster 2 until they were discharged in the final week of the study, and cluster 2 persisted in the room 7 environment until the end of the study. Persistence of cluster 2 in room 7 was associated with a second patient acquisition event, by patient 127 a week after they were admitted to the ICU and confirmed CRAB-negative. Movement of cluster 2 to rooms 5 and 11 appeared to be driven by the movement of patient 97 (who produced cluster 2 until the end of the study). Similarly, bed unit 17 in room 7 was contaminated with CRAB cluster 2 isolates. The patient occupying this bed, P121, was not found to be colonised by cluster 2 but their syringe driver was found to be contaminated after they moved to bed unit 7 in room 4. Cluster 2 was also found on another syringe driver in bed unit 23 in room 10. This may have been associated with ICU staff who would be most likely to touch this instrument.

**Figure 4:**
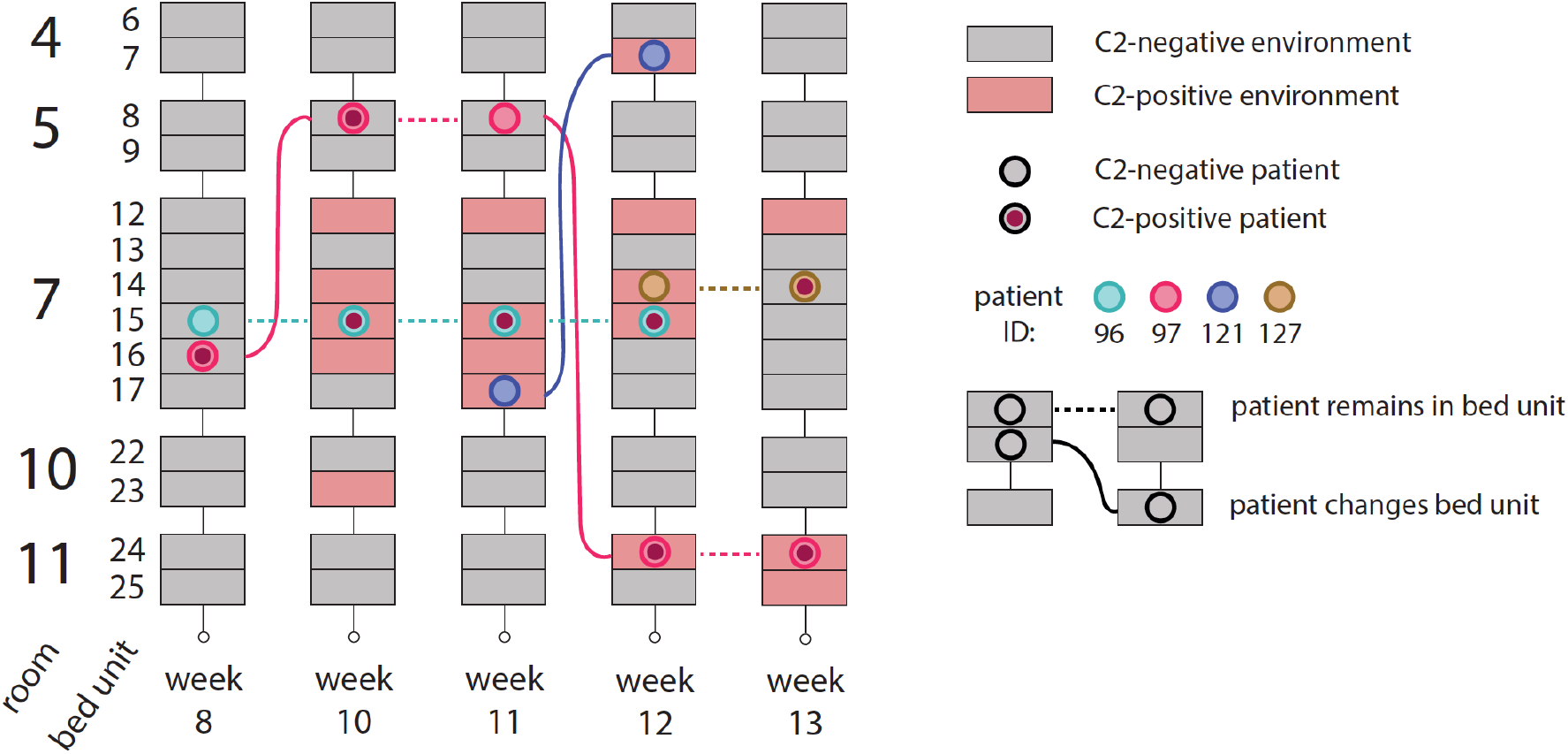
Spatiotemporal distribution of cluster 2 isolates in the ICU. Schematic showing the distribution of all GC2 cluster 2 (C2) isolates in rooms 4, 5, 7, 10 and 11 over weeks 8 to 13 of the study. These were the only instances where C2 was isolated over the course of the study. Sampling dates (day/month) are shown in brackets. Each bed unit is shown as a rectangle, shaded red when C2 was isolated from the environment and grey when it was not. Patients are shown as coloured circles, with a red fill indicating the presence of C2 in patient-screening samples. Dashed horizontal lines represent a patient’s continued presence in a bed unit while solid lines show the relocation of patients between sampling dates.

The spatiotemporal distribution of clusters was used to assess potential patterns of CRAB movement and persistence in the ICU (Figure 5). Of 119 occasions when rooms were found to contain CRAB, the same cluster was found in multiple bed units 46 times (38.66 %) suggesting it may have spread between them. On 70/119 occasions (58.82 %), rooms adjacent to one another had the same CRAB cluster suggesting that strain movement had potentially occurred, though the number or direction of events could not be determined (Table S7) and re-introduction of the cluster could be responsible for the CRAB distribution. Of the 116 occasions when complete sample sets were taken from different consecutive occupants of a single bed unit, the same CRAB clusters were found associated with consecutive patients on 16 occasions (13.79 %; Table S8). The majority of potential strain movement between beds, rooms and consecutive patients could be solely accounted for by looking at environmental contamination (Tables S7 and S9). Six patients that were CRAB-negative within two days of admission later yielded CRAB-positive oral or rectal swabs. The CRAB clusters in these oral or rectal swabs had been present in their respective patients’ bed unit or room environments before the patients were admitted. These cases provide clear evidence for patient acquisition of CRAB from contaminated ICU environments.

**Figure 5:**
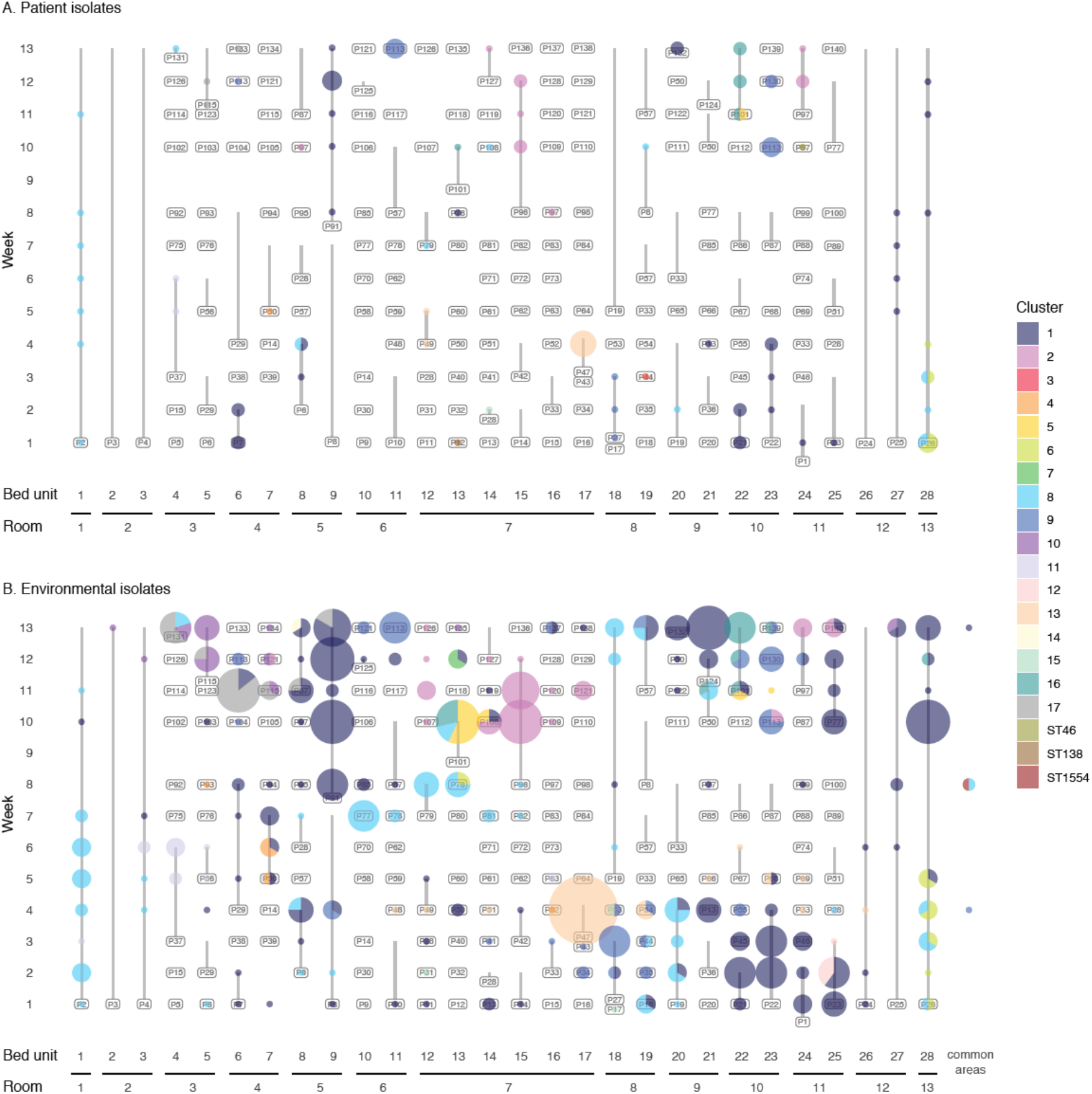
Diversity and distribution of CRAB isolates within the ICU. Distribution of isolates from each CRAB cluster; bed unit numbers are indicated across the x-axis and reflect the spatial arrangement of beds in the ICU (Figure S1); isolates from common areas are grouped together but may be from spatially distinct locations; the vertical axis shows the week of sampling; labels show the ID of the patient occupying the bed with vertical lines extending from the date of first to last sample associated with the patient; size of coloured bubbles reflects the number of CRAB isolates found in the patient and their bed unit on each date of sampling; the colour of bubbles corresponds to the CRAB cluster; multi-coloured bubbles are split as a pie chart to reflect the proportion of isolates from each cluster. A) Isolates from all samples, B) Isolates from patient screening and clinical samples only, B) Isolates from environmental samples only.

### Plasmids were transmitted between CRAB clusters during the study

Using the pAci database created for this study we detected eight plasmid replicon types in the CRAB collection (Table S10). Aci9 (448 isolates), Aci6 (126 isolates), Aci1 (75 isolates), Aci2a (19 isolates), and GR24-type replicons were found amongst GC2/ST187 clusters (Figure 1). A 38,952 bp plasmid, pDETAB14, was found only in the cluster 11 isolate DETAB-E154 and represents a novel type (Figure 1; Table S10). The distributions of Aci9, Aci1, and Aci2a replicons were largely consistent with the GC2/ST187 phylogeny (Figure 1) and complete sequences showed that they were associated with well-conserved small plasmids pDETAB9, pDETAB10, and pDETAB11, respectively (Table S10). Identical Aci6 *rep* genes were found in multiple, disparate clusters (Figure 1) and a single Aci6 *rep* gene with one SNP relative to the others was found in isolate DETAB-P90.

Three plasmid backbone types, represented by pDETAB7, pDETAB8, and pDETAB16, were identified amongst complete Aci6 plasmid sequences (Figure S3; Table S10). Four variants of pDETAB7 were distinguished by insertions or SNPs in otherwise identical 72 kbp backbones – pDETAB7a (no insertions), pDETAB7b (IS*Aba1* insertion), pDETAB7c (Tn*6022* insertion), and pDETAB7d (Tn*6022* insertion and two backbone SNPs) (Figure S3). The conjugative transfer regions of all Aci6 plasmids found here were complete and uninterrupted. The distinct Aci6 plasmids were detected in draft genomes using the unique sequences of their characteristic insertions or backbone regions (Table S11), revealing their distributions amongst the CRAB population (Figure 6). The Aci6 plasmids were found in one to six CRAB clusters each. The heterogeneous presence of plasmids in polyphyletic clusters strongly suggested that they had transferred recently, possibly facilitated by the extensive co-location of diverse clusters observed throughout the ICU.

**Figure 6.**
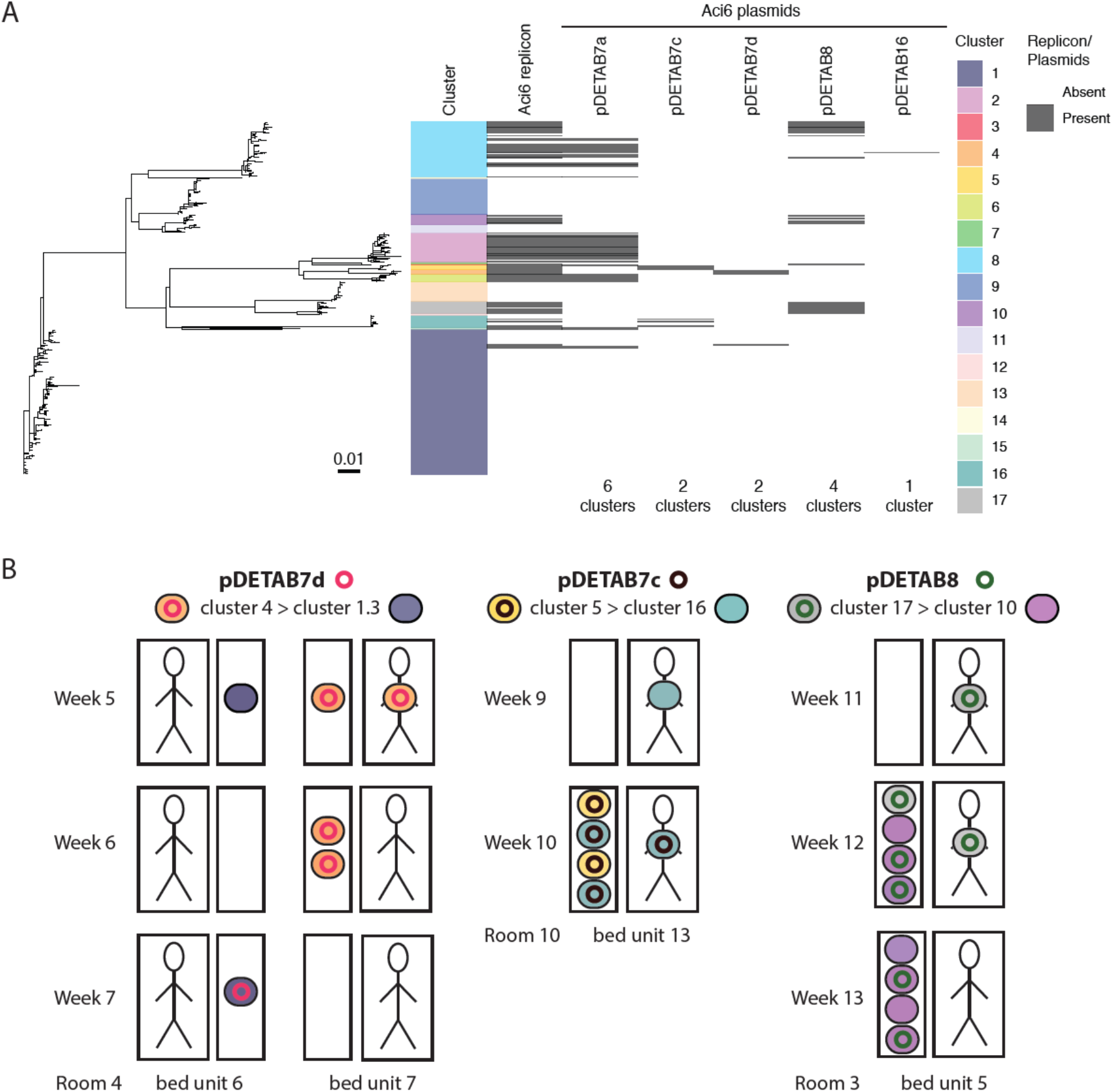
Dissemination of Aci6 plasmids amongst CRAB clusters. A) Phylogenetic relationships of all GC2/ST187 isolates; tracks adjacent to the tips show isolate cluster designations, presence of the Aci6 plasmid replicon detected by pAci, and the presence of specific Aci6 plasmids determined from hybrid genome assemblies or detection of signature sequences in draft genomes. B) Schematic overview of putative pDETAB7d, pDETAB7c and pDETAB8 transfer events. Bed units are shown as paired boxes that represent patient-derived (larger boxes containing figurative person) and environmental (small boxes) sources of isolation. Weeks of isolation and room/bed unit numbers are labelled. Isolates are shown as coloured ovals and plasmids as coloured circles. The identities of plasmids and isolate clusters, and the direction or putative plasmid transfer are indicated at the top of the panel. Only isolates of cluster 1 subcluster 3 thought to be involved in the transfer event are shown.

To explore the possibility of Aci6 plasmid transfer in the ICU, we searched for instances where isolates of a given cluster carrying a particular plasmid had been isolated after or at the same time as isolates of the same cluster that lacked the plasmid. In such cases, we checked whether isolates of a different cluster bearing the same plasmid had been present in the same location at the same time. We defined putative transfer events as situations where, for a given plasmid, a plasmid-containing isolate of one cluster (“donor”) was present in the same room within two weeks of plasmid-free (“recipient”) and plasmid-containing (“transconjugant”) isolates of a different cluster. Three putative transfer events were identified, involving three different Aci6 plasmids, six different clusters and three different rooms (Figure 6B). In all cases we confirmed that the core genomes of putative recipients and transconjugants were identical and compared contigs extracted from the draft genomes of donors and transconjugants to confirm that at least 50 kbp of their Aci6 plasmid backbones were identical. Thus, we conclude that three different Aci6 plasmids transferred between clusters in the ICU over the course of this study.

## Discussion

This study assessed the CRAB population found in an ICU’s environment, staff and patients. It has provided insights into the abundance and extensive diversity of CRAB found in a single ICU population, the dynamics of cluster introductions to the ICU, dissemination within the ICU, and the dissemination of *Acinetobacter* plasmids in a hospital setting. Our deep-sampling approach revealed previously unappreciated diversity in the ICU’s endemic predominantly GC2 CRAB population (Fig 1). This situation, in a country where CRAB prevalence is reported to be highest globally,^28^ is in stark contrast to those described in monoclonal outbreaks in hospitals in low-prevalence high-income countries.^4,7–9^ The prevalence of CRAB in the ICU was also found to be higher than previously appreciated, likely owing to our environmental screening.

Analysis indicated that there was a large reservoir of CRAB in the hospital environment that was introduced to the ICU on multiple occasions in association with patient admissions. Only 53 patients were swabbed within two days of admission, so patient-associated importation was likely underestimated. The ICU environment was heavily contaminated with CRAB, given that a bed unit yielded CRAB on 183/335 (54.63 %) sampling occasions, relative to 72/299 (24.08 %) occasions when patients were sampled. This environmental contamination appeared to drive dissemination within the ICU and a CRAB cluster in one room was often found in multiple bed units and adjacent rooms. Of 35 CRAB-positive patients in the study, 14 acquired CRAB during their ICU stay, and the acquired clusters were often found in the patient’s immediate environment. Together these results highlight the important role of the environment in CRAB persistence and eventual acquisition by patients, and the need to target this reservoir with IPC measures. Our data suggests these might include: regular and appropriate cleaning, particularly of patient rooms and mobile equipment, isolation of patients known to carry CRAB, minimisation of patient relocation between beds, and enhanced hand-washing and decontamination procedures, as many surfaces that yielded CRAB are only touched by staff.

Routine diagnostic screening over the course of this study isolated just 19 CRAB isolates despite extensive contamination of the environment and colonisation of patients. Only considering clinical samples, as is done in other studies on CRAB diversity among hospitalised patients,^13,14,29^ would clearly have led to a vast underestimate of the CRAB population in this ICU. It is clear from this study that without considering patient screening and environmental isolates it will be near-impossible to accurately assess complex polyclonal CRAB transmission pathways associated with infections. We postulate that environmental surveillance in areas shown to be frequently contaminated and rigorous screening of patients would present a more sensitive strategy for monitoring wards and beginning to reduce the CRAB burden in such settings.

Co-localisation of distinct CRAB clusters in a single bed unit was a common occurrence in this setting. This was observed despite examination of just a single isolate from each CRAB-positive sample, and suggests that further cluster co-localisation or persistence might have gone undetected. Extensive co-localisation facilitated three putative Aci6 plasmid transfer events that were identified over the course of the study. Aci6 plasmids are known to carry antibiotic resistance genes, including carbapenemase genes,^30–32^ although the plasmids involved in transfer events here did not. In the laboratory, Aci6 plasmids have been shown to be conjugative and to mediate the mobilisation of small antibiotic resistance plasmids.^32^ Our observations here provide the first evidence that Aci6 plasmids transfer readily under hospital conditions in the absence of an obvious fitness advantage. The provision of conditions suitable for horizontal gene transfer is concerning, and our findings suggest that the introduction of plasmid-borne antibiotic resistance determinants might result in their rapid dissemination through hospital populations. WGS efforts employed in concert with infection prevention and control measures targeting AMR transmission must consider plasmids and other mobile genetic elements,^33^ and the implementation of our pAci database will facilitate the examination of *Acinetobacter* plasmids in future studies.

### Data sharing

Raw sequence data for all isolates is available via National Center for Biotechnology Information under BioProject accession PRJNA738868.

## Acknowledgements

We thank Prof. Zhiyong Zong and his team at West China Hospital for their careful teaching of sampling methods and are grateful to the doctors and nurses in the ICU for performing patient sampling. We appreciate the use of servers provided by MRC CLIMB BIG DATA grant MR/T030062/1.

