## Supplementary methods for "Extensive acquisition of carbapenem-resistant *Acinetobacter baumannii* in Intensive Care Unit patients is driven by widespread environmental contamination"

**Planned sampling sites**

●	**Weekly sampling sites:** Bed rails, ventilators, ECG monitors, syringe drivers, switch buttons, nebulisers, stethoscopes, ventilator shelves, bed controllers, bedside tables, computers, infusion stands, dispensing trolleys and lockers

●	**Fortnightly sampling sites:** sink sites in the bed units: Inside sink drains, tap surfaces, sink countertops and inner-wall of overflows

●	**Monthly sampling sites:**

○	Nurse station: Computers, phones, barcode printer, barcode scanner, inside sink drains, tap surfaces, inner wall of basins, sink countertops, the inner wall of overflows and water in sink trap;

○	Room 10 and 7: crash trolleys

○	Corridor: UV disinfector, Ozone disinfector, sputum suction machines, non-invasive ventilators

○	Storage room: Switch buttons, instrument cabinets

○	Bathroom: Door handle 1 and 2

○	Cleaning room: Cleaning trolley cover, cleaning trolley handle, cleaning trolley body and mop handle

Staff working in the ward participated in the study voluntarily, providing rectal swabs or stool samples at the beginning of every month. Patient screening samples were planned to be collected within two days of patient admission then weekly thereafter from oral swabs and rectal swabs, and when present, the nasogastric tube, nasojejunal tube, endotracheal tube and tracheostomy tube of patients. CRAB isolated from samples sent to the clinical lab were also collected. Spatiotemporal details of sample collection and relevant patient information was additionally collated.

**Sample collection**

Environmental surfaces (approximately 10x10 cm) were swabbed with Sterile swabs (COPAN, Italy) pre-moistened with 2 mL tryptone soy broth (TSB, Oxoid, Hampshire, UK) supplemented with 0·1% sodium thiosulfate (Sangon Biotech, China). The swabs were immediately placed into 14 ml sterile tubes containing 2mL TSB with 0·1% sodium thiosulfate then incubated for 24 hours at 37°C. After culture, 20 µL overnight bacteria were streaked onto to Acinetobacter CHROMagar plates (CHROMagar, Paris, France) containing 2µg/mL meropenem followed by overnight incubation at 37°C.

For patient and staff samples, rectal swabs containing Cary-Blair Transport Medium (Gongdong, Taizhou, China) were used and other swabs were moistened using 0·9% saline before sampling. These swabs were plated directly onto Acinetobacter CHROMagar plates supplemented with 2 µg/mL meropenem and again incubated at 37°C for 24 hours. After subculture, a single, isolated colony of presumptive *A. baumannii* was selected based on red colour and morphology, streaked onto a Mueller-Hinton agar plate (Oxoid, Hampshire, UK) and incubated at 37°C for 24 hours. A single, isolated colony from the MH plate was collected and then identified as *Acinetobacter baumanii* by Matrix-assisted laser desorption ionization-time of flight mass spectrometry (MALDI-TOF MS) (bioMérieux, France) and 16S rRNA gene sequencing using the primers 27F-5’- AGAGTTTGATCCTGGCTCAG -3’ and 1492R-5’- GGTTACCTTGTTACGACTT -3’. Carbapenem resistance was confirmed using minimum inhibitory concentrations for imipenem and meropenem, determined using agar microdilution with results interpreted according to the Clinical and Laboratory Standards Institute 2019 guidelines; Escherichia coli ATCC 25922 served as the quality control strain.

**Illumina sequencing**

DNA was prepared for Illumina sequencing from the confirmed CRAB isolates. One colony from each purified culture was cultured in 2 mL Mueller-Hinton broth for 24 hours at 37°C. Cell pellets were harvested and added to 180 µL Buffer ATL with 20 µL Proteinase K (Qiagen, Germany), incubating for 2 h. Then, 200 µL Buffer AL was added and the mixture was incubated at 70°C for 10 minutes. The mixture was transferred into the QIAamp Mini spin column and centrifuged at 12,000 × g for 30 seconds. Next, the column was washed using Buffer AW1 and AW2. DNA was eluted using distilled water by centrifugation at 10,000 × g for 30 seconds. The quality and quantity of DNA were assessed using a NanoDrop 2000 (Thermo Scientific, USA) and Qubit 4.0 fluorometer (Invitrogen, USA).

Illumina sequencing libraries were prepared using the TruePrepTM DNA Library Prep Kit V2 (Vazyme) according to the manufacturer’s protocol. Individual libraries were assessed on the QIAxcel Advanced Automatic nucleic acid analyzer using a high-resolution gel cartridge (Qiagen, Germany), and then were quantified by qPCR using the use of KAPA SYBR FAST qPCR Kits Kapa KK4610 (KAPA Biosystem, Wilmington, MA, U.S.A.). Paired-end sequencing (2×150-bp reads) was performed on the Illumina HiSeq X Ten platform (Illumina Inc., San Diego, CA, USA).

**Oxford Nanopore sequencing**

Long-read sequencing was undertaken for 60 isolates selected after Illumina sequencing on the basis of phylogenetic, resistance gene and plasmid diversity. For this, DNA was extracted using the Gentra® Puregene® Yeast/Bact. Kit (Qiagen, Germany) according to manufacturer’s protocol with minor modifications. Briefly, cell pellets were harvested into a sterile 15 mL falcon tube from 5mL overnight culture. Each pellet was resuspended in 500 µL Lysis Buffer Solution, transferred to a Phase Lock tube and incubated at 37°C for 30 minutes. Cell lysates were treated with 2 µL RNase A (100 mg/mL, Qiagen, Germany), gently mixed by flicking the tube and incubated at 37°C for 30 minutes. The cell lysate was further treated with 3 µL Proteinase K (Qiagen, Germany), mixed by gentle inversion and incubated at 50°C for 90 minutes. The suspension was mixed with 500 µL phenol:chloroform:isoamyl alcohol (25:24:1) by inversion and centrifuged at 20,000 rcf for 10 minutes. The supernatant was collected and DNA was purified using AMPure XP beads (Beckman Coulter™) with minor modifications to the manufacturer’s protocol: Fresh 70% ethanol was used for washing the DNA on the beads and after ethanol was removed with a pipette, the beads were incubated on a 37°C heat block for 5 min to dry off residual ethanol. The DNA was resuspended in 150 µl of nuclease-free water. The quality and quantity of DNA were then assessed using a NanoDrop 2000 (Thermo Scientific, USA) and Qubit 4.0 fluorometer (Invitrogen, USA) with the dsDNA BR assay kit (Thermo Scientific, USA).

Oxford Nanopore sequencing libraries were prepared using the SQU-LSK109 Ligation Sequencing kit (Oxford Nanopore Technologies, UK) in conjunction with the PCR-Free ONT EXP-NBD104 Native Barcode Expansion kit (Oxford Nanopore Technologies, UK) according to the native barcoding genomic DNA protocol. DNA was processed without the optional shearing steps to select for long reads. After quantification of the individual libraries by the Qubit and normalisation of library concentrations, the library was sequenced on the GridION X5 platform (Oxford Nanopore Technologies, UK).

**Bioinformatic analyses**

Illumina sequence reads were trimmed and assembled with Shovill v1.1.0 under default settings but with a 10x minimum contig coverage (https://github.com/tseemann/shovill). Read quality was determined with FastQC v0.11.8,^1^ and assemblies were assessed for contamination and completeness using Centrifuge v1.0.4, QUAST v5.0.2, CheckM v1.0.13 and ARIBA v2.14.1 to identify heterogeneity in MLST genes.^2–4^ All genomes meeting quality expectations had a total genome size of 3,800,935 - 4,150,853bp; N50 ≥ 27,912; GC content of 38·77 – 39·1 %; genome completeness of ≥99·62%; ≤0·5% contamination; ≤288 contigs and complete MLST genes without nucleotide heterogeneity. For hybrid assemblies, Nanopore reads were trimmed with Filtlong v0.2.0 (https://github.com/rrwick/Filtlong) under default settings targeting approximately 100-fold genome coverage. These were assembled with the trimmed Illumina reads using Unicycler v0.4.8.^5^

Phylogenies were generated using the following reference genomes: DETAB-E107 for the GC2/ST187 population; DETAB-E47 as an outgroup for cluster 1 isolates; DETAB-H50 internal reference for cluster 1 isolates and DETAB-P90 for cluster 8. All other bioinformatic analyses were conducted as described in the manuscript with the additional detail that Gubbins was run using settings that built phylogenies over five iterations using RaxML and a GTRGAMMA model.

**Data visualisation and interpretation**

Bioinformatic outputs and metadata were analysed in R,^6^ using tidyverse version 1.3.0 packages.^7^ Visualisation of phylogenetic trees (fig. 1 and fig. 3a) also used ggtree version 2.0.1 and ggnewscale version 0.4.5.^8,9^ Creation of “Bubble diagrams” (fig. 2b and fig. S1) used the additional scatterpie version 0.1.5,^10^ ggthemes version 4.2.0,^11^ and scales version 1.1.1 packages.^12^ The minimum spanning tree (fig. 2a) was produced with GrapeTree from the RAxML tree.^13^ Editing of figures and production of the schematic diagram (fig. 3b) was undertaken in Adobe Illustrator.^14^

9.	Campitelli E. ggnewscale: Multiple Fill and Colour Scales in ‘ggplot2’. Available from: https://CRAN.R-project.org/package=ggnewscale

10.	Yu G. scatterpie: Scatter Pie Plot. Available from: https://CRAN.R-project.org/package=scatterpie

11.	Arnold JB, Daroczi G, Werth B, Weitzner B, Kunst J, Auguie B, et al. ggthemes: Extra Themes, Scales and Geoms for ‘ggplot2’. Available from: https://CRAN.R-project.org/package=ggthemes

12.	Wickham H, Seidel D, RStudio. scales: Scale Functions for Visualization. 2020. Available from: https://CRAN.R-project.org/package=scales

13.	Zhou Z, Alikhan N-F, Sergeant MJ, Luhmann N, Vaz C, Francisco AP, et al. GrapeTree: Visualization of core genomic relationships among 100,000 bacterial pathogens. Genome Res. 2018 Jul;gr.232397.117.

14.	Industry-leading vector graphics software | Adobe Illustrator. Available from: https://www.adobe.com/products/illustrator.html
