## supplementary tables for "Extensive acquisition of carbapenem-resistant *Acinetobacter baumannii* in Intensive Care Unit patients is driven by widespread environmental contamination"

**Table S1: Sequences in the pAci *Acinetobacter* plasmid typing database**

| **Plasmid type** | **Rep**  **Group** | **Reference plasmid** | | **Notes** |
| --- | --- | --- | --- | --- |
|  |  | **Name** | **Accession** |  |
| GR1 | GR1 | p1ABSDF | CU468231 | ·· |
| Aci1 | GR2 | pACICU1 | CP031381 | ·· |
| Aci3 | GR3 | p203 | GU978997^4^ | CP072529 is complete |
| Aci7 | GR3 | p736 | GU978996 | no complete plasmid in GenBank |
| pS30-1 | GR3 | pS30-1 | KY617771 | ·· |
| Aci4 | GR4 | p844 | GU978998^4^ | CP040045 is complete |
| Aci5 | GR5 | p537 | GU978999^4^ | CP044480 is complete (5-SNP *rep*) |
| Aci6 | GR6 | pACICU2 | CP031382 | ·· |
| GR7 | GR7 | p3ABSDF | CU468233 | ·· |
| pXBB1-2 | GR7 | pXBB1-2 | CP010353 | ·· |
| Aci8 | GR8 | p11921 | GU979000^4^ | KY984087 is complete |
| Aci9 | GR8 | pMAC | AY541809 | ·· |
| repA1343 | GR8 | pA1343 | MN461226 | ·· |
| GR9 | GR9 | p3ABSDF | CU468233 | ·· |
| AciX | GR10 | pACICU1 | CP031381 | ·· |
| GR11 | GR11 | p1ABAYE | CU459137 | ·· |
| repA_AB | GR12 | pAB02 | AY228470^4^ | GQ342610 is complete |
| pABIR | GR12 | pABIR | EU294228 | ·· |
| GR13 | GR13 | p3ABAYE | CU459140 | ·· |
| AciRCR1 | GR14 | p4ABAYE | CU459139 | ·· |
| GR15 | GR15 | p3ABSDF | CU468233 | ·· |
| AciRCR2 | GR16 | pAB49 | L77992^4^ | CP030108 is complete (1-SNP *rep*) |
| AciRCR4 | GR16 | pA85-1a | CP021784 | ·· |
| GR17 | GR17 | pAB1 | CP000522 | ·· |
| GR18 | GR18 | p2ABSDF | CU468232 | ·· |
| GR19 | GR19 | p135040 | GQ861437 | ·· |
| Aci2a | GR20 | pABVA01 | FM210331 | ·· |
| Aci2b | GR20 | pD72-1 | KM051986 | ·· |
| GR21 | GR21 | pAba242_12 | KY984046 | ·· |
| GR22 | GR22 | pAba242_25 | KY984047 | ·· |
| GR24 | GR24 | pABTJ2 | CP004359 | ·· |
| GR25 | GR25 | pABTJ1 | CP003501 | ·· |
| GR26 | GR26 | pAba3207a | CP015365 | ·· |
| GR27 | GR27 | pABUH2b-5.4 | AYFH01000057^4^ | CP045125 is complete |
| GR28 | GR28 | pNaval81-26 | AFDB02000003 | ·· |
| GR29 | GR29 | pAba9201a | CP023021 | ·· |
| GR30 | GR30 | pABUH5-114 | AYOI01000002^4^ | CP026944 is complete |
| GR31 | GR31 | pD36-4 | CP012956 | ·· |
| GR32 | GR32 | pKBN10P02143 | CP013925 | ·· |
| GR33 | GR33 | pAB3 | CP012005 | ·· |
| GR34 | GR34 | pDETAB2 | CP047975 | ·· |
| AciRCR3 | GR35^1^ | p3AB5075 | CP008709 | ·· |
| pMS32-3 | GR36^1^ | pMS32-3 | KJ616406 | ·· |
| repA352 | GR37^1^ | pA352 | MN461227 | ·· |
| repA2949 | GR38^1^ | pA2949 | MN481287 | ·· |
| pRAY | NT^2^ | pRAY* | JQ904627 | targets *mobAC* mobilisation genes |
| pA297-3 | NT^2^ | pA297-3 | KU744946 | targets putative partitioning gene |
| pRCH51-3 | NT^2^ | pRCH51-3 | KY216144 | targets putative partitioning gene |
| pDETAB14 | NT^2^ | pDETAB14 | CP077844 | targets putative relaxase gene |
| pSU1904NDM | NT^2^ | pSU1904NDM | LC537594 | targets putative relaxase gene |
| pR4WN | NT^2^ | pR4WN_12CE1 | MT742180 | targets putative partitioning gene |
| *repAciN* | -^3^ | AbGRI3 | KX011025.2 | *rep*-like fragment in AbGRI3 |

^1^ Rep proteins encoded by these plasmids are <74% identical to the Rep proteins of existing groups, so were assigned new names here. Note that a representative for group GR23 does not appear in the pAci database because the sequence that was named GR23 (PMID: 32625185) is identical to the Aci8/GR8 sequence.

^2^ NT = not typeable. These plasmids do not contain a recognisable *rep* gene and therefore cannot be assigned a to a Rep group. In the absence of *rep* genes, alternative backbone sequences have been used to represent these plasmid types in pAci. The identities of target sequences are listed in the ‘Notes’ column.

^3^ The *repAciN* gene found in the chromosomal resistance island AbGRI3 is included here to prevent false identifications of AbGRI3 as an Aci7-like plasmid. AbGRI3 is not a plasmid.

^4^ This accession does not contain a complete plasmid sequence. The ‘Notes’ column lists alternative GenBank entries that contain complete plasmids with identical or almost identical *rep* genes (number of SNPs relative to the reference *rep* gene shown in brackets where different).

**Table S2: Summary of samples and isolates by sample type**

Count of each sample type collected and the CRAB isolates yielded with percentage of samples yielding CRAB; count of distinct patients sampled or associated with the bed unit environment sampled and those yielding isolates, with percentage of those sampled yielding CRAB. Note, samples from bed units with no patient present are excluded from counts of distinct patients but included in counts of total samples and isolates yielded.

| **Collection type** | **Sample type** | **Room type** | **# samples** | **# isolates** | **% Positive samples** | **# distinct associated patients sampled** | **# distinct associated patients yielding CRAB** | **% Positive associated patients** |
| --- | --- | --- | --- | --- | --- | --- | --- | --- |
| Clinical sample | Ascitic fluid | Bed unit | 1 | 1 | 100·00 | 1 | 1 | 100·00 |
| Clinical sample | Bile | Bed unit | 1 | 1 | 100·00 | 1 | 1 | 100·00 |
| Clinical sample | Hydrothorax | Bed unit | 1 | 1 | 100·00 | 1 | 1 | 100·00 |
| Clinical sample | Pelvic drainage fluid | Bed unit | 1 | 1 | 100·00 | 1 | 1 | 100·00 |
| Clinical sample | Sputum | Bed unit | 15 | 15 | 100·00 | 11 | 11 | 100·00 |
| Patient screening | Rectal swab | Bed unit | 289 | 41 | 14·19 | 118 | 12 | 10·17 |
| Patient screening | Oral swab | Bed unit | 254 | 32 | 12·60 | 112 | 14 | 12·50 |
| Patient screening | Nasogastric tube | Bed unit | 169 | 15 | 8·88 | 68 | 6 | 8·82 |
| Patient screening | Nasojejunal tube | Bed unit | 37 | 2 | 5·41 | 25 | 2 | 8·00 |
| Patient screening | Tracheostomy tube | Bed unit | 143 | 7 | 4·90 | 33 | 3 | 9·09 |
| Patient screening | Endotracheal tube | Bed unit | 72 | 3 | 4·17 | 49 | 1 | 2·04 |
| Environment | Ventilator | Bed unit | 287 | 80 | 27·87 | 111 | 19 | 17·12 |
| Environment | Dispensing trolley | Bed unit | 39 | 10 | 25·64 | 21 | 4 | 19·05 |
| Environment | Ventilator shelf | Bed unit | 278 | 41 | 14·75 | 109 | 18 | 16·51 |
| Environment | Nebuliser | Bed unit | 281 | 39 | 13·88 | 110 | 17 | 15·45 |
| Environment | Syringe driver | Bed unit | 279 | 37 | 13·26 | 130 | 14 | 10·77 |
| Environment | Bed rail | Bed unit | 332 | 44 | 13·25 | 136 | 9 | 6·62 |
| Environment | Bedside table | Bed unit | 335 | 40 | 11·94 | 137 | 18 | 13·14 |
| Environment | Sink countertop | Bed unit | 52 | 6 | 11·54 | 28 | 3 | 10·71 |
| Environment | ECG monitor | Bed unit | 333 | 34 | 10·21 | 137 | 11 | 8·03 |
| Environment | Infusion stand | Bed unit | 335 | 31 | 9·25 | 137 | 13 | 9·49 |
| Environment | Inside sink drain | Bed unit | 52 | 4 | 7·69 | 28 | 4 | 14·29 |
| Environment | Computer | Bed unit | 194 | 12 | 6·19 | 95 | 6 | 6·32 |
| Environment | Bed controller | Bed unit | 332 | 20 | 6·02 | 136 | 8 | 5·88 |
| Environment | Stethoscope | Bed unit | 315 | 18 | 5·71 | 133 | 11 | 8·27 |
| Environment | Locker | Bed unit | 167 | 8 | 4·79 | 72 | 3 | 4·17 |
| Environment | Tap surface | Bed unit | 52 | 1 | 1·92 | 28 | 0 | 0·00 |
| Environment | Switch button | Bed unit | 191 | 3 | 1·57 | 82 | 2 | 2·44 |
| Environment | Inner wall of overflow | Bed unit | 51 | 0 | 0·00 | 27 | 0 | 0·00 |
| Environment | Crash trolley 2 | Bedroom | 2 | 1 | 50·00 | NA | NA | NA |
| Environment | Crash trolley 1 | Bedroom | 3 | 1 | 33·33 | NA | NA | NA |
| Environment | Door handle1 | Bathroom | 3 | 0 | 0·00 | NA | NA | NA |
| Environment | Door handle2 | Bathroom | 3 | 0 | 0·00 | NA | NA | NA |
| Environment | Cleaning trolley cover | Cleaning room | 3 | 1 | 33·33 | NA | NA | NA |
| Environment | Cleaning trolley body | Cleaning room | 3 | 0 | 0·00 | NA | NA | NA |
| Environment | Cleaning trolley handle | Cleaning room | 3 | 0 | 0·00 | NA | NA | NA |
| Environment | Mop handle | Cleaning room | 3 | 0 | 0·00 | NA | NA | NA |
| Environment | Non-invasive ventilator 1 | Corridor | 1 | 0 | 0·00 | NA | NA | NA |
| Environment | Non-invasive ventilator 2 | Corridor | 1 | 0 | 0·00 | NA | NA | NA |
| Environment | Ozone disinfector | Corridor | 1 | 0 | 0·00 | NA | NA | NA |
| Environment | Sputum suction 1 | Corridor | 3 | 0 | 0·00 | NA | NA | NA |
| Environment | Sputum suction 2 | Corridor | 2 | 0 | 0·00 | NA | NA | NA |
| Environment | UV disinfector | Corridor | 1 | 0 | 0·00 | NA | NA | NA |
| Environment | Inner wall of basin | Nurse station | 3 | 1 | 33·33 | NA | NA | NA |
| Environment | Barcode printer | Nurse station | 3 | 0 | 0·00 | NA | NA | NA |
| Environment | Barcode scanner | Nurse station | 3 | 0 | 0·00 | NA | NA | NA |
| Environment | Computer 1 | Nurse station | 3 | 0 | 0·00 | NA | NA | NA |
| Environment | Computer 2 | Nurse station | 3 | 0 | 0·00 | NA | NA | NA |
| Environment | Computer 3 | Nurse station | 3 | 0 | 0·00 | NA | NA | NA |
| Environment | Inner wall of overflow | Nurse station | 2 | 0 | 0·00 | NA | NA | NA |
| Environment | Inside sink drain | Nurse station | 3 | 0 | 0·00 | NA | NA | NA |
| Environment | Phone 1 | Nurse station | 3 | 0 | 0·00 | NA | NA | NA |
| Environment | Phone 2 | Nurse station | 2 | 0 | 0·00 | NA | NA | NA |
| Environment | Sink countertop | Nurse station | 3 | 0 | 0·00 | NA | NA | NA |
| Environment | Tap surface | Nurse station | 3 | 0 | 0·00 | NA | NA | NA |
| Environment | Water in sink trap | Nurse station | 2 | 0 | 0·00 | NA | NA | NA |
| Environment | Instrument cabinet 1 | Storage room | 3 | 0 | 0·00 | NA | NA | NA |
| Environment | Instrument cabinet 2 | Storage room | 3 | 0 | 0·00 | NA | NA | NA |
| Environment | Instrument cabinet 3 | Storage room | 3 | 0 | 0·00 | NA | NA | NA |
| Environment | Switch button | Storage room | 3 | 0 | 0·00 | NA | NA | NA |

ECG echocardiogram; UV ultraviolet; NA Not applicable

**Table S3: SNP distance data in each of the identified CRAB clusters**

Table shows within cluster SNP distances and distance to nearest cluster based on data from SNIPPY mapping as outlined in supplementary methods.

|  | Next-closest cluster | | Within-cluster SNP-dists | | | | | |  |
| --- | --- | --- | --- | --- | --- | --- | --- | --- | --- |
| Cluster | Cluster | Minimum  SNP-dist | Min. | 1^st^ Qu. | Median | Mean | 3^rd^ Qu. | Max. | IQTree-predicted  MRCA |
| 1 | 11 | 57 | 0 | 2 | 4 | 3.92 | 5 | 24 | 09/07/2019 |
| 2 | 7 | 48 | 0 | 5 | 8 | 7.84 | 10 | 26 | 28/05/2017 |
| 3 | 2 | 52 | NA | NA | NA | NA | NA | N/A | 20/08/2019 |
| 4 | 5 | 35 | 0 | 1 | 4 | 2.87 | 4 | 8 | 27/08/2019 |
| 5 | 4 | 35 | 0 | 0 | 1 | 1.32 | 2 | 5 | 03/03/2019 |
| 6 | 2 | 62 | 0 | 3.25 | 5 | 5.27 | 7 | 11 | 01/03/2019 |
| 7 | 2 | 48 | 0 | 0 | 0 | 0 | 0 | 0 | 14/07/2019 |
| 8 | 11 | 71 | 0 | 2 | 5 | 5.27 | 8 | 17 | 07/04/2019 |
| 9 | 11 | 33 | 0 | 1 | 2 | 2.03 | 3 | 11 | 31/10/2018 |
| 10 | 11 | 13 | 0 | 1 | 2 | 2.1 | 3 | 8 | 12/05/2019 |
| 11 | 10 | 12 | 0 | 2 | 3 | 5.77 | 8 | 20 | 2019 |
| 12 | 13 | 64 | 0 | 0.5 | 1 | 0.667 | 1 | 1 | 04/06/2019 |
| 13 | 17 | 62 | 0 | 0 | 1 | 0.852 | 1 | 3 | 10/08/2019 |
| 14 | 11 | 72 | 2 | 2 | 2 | 2 | 2 | 2 | 08/10/2019 |
| 15 | 11 | 86 | 1 | 69 | 76.5 | 70.2 | 94.5 | 100 | 12/06/2004 |
| 16 | 15 | 117 | 0 | 0 | 1 | 1.29 | 2 | 4 | 21/08/2019 |
| 17 | 12 | 18 | 0 | 2 | 3 | 3.86 | 5 | 12 | 14/03/2019 |
| ST138 | - | - | - | - | - | - | - | - | - |
| ST1554 | - | - | - | - | - | - | - | - | - |
| ST46 | - | - | - | - | - | - | - | - | - |

**Table S4: MRCAs for each GC2/ST187 cluster**

Predicted dates of most recent common ancestor (MRCA) for isolates within each cluster and between the given cluster and any other cluster identified in the ICU.

| **Cluster** | **Predicted date of MRCA for all isolates within cluster** | **Predicted date of MRCA with any other cluster** |
| --- | --- | --- |
| 1 | 09/07/19 | 18/04/04 |
| 2 | 28/05/17 | 15/05/10 |
| 3 | 20/08/19 | 15/05/10 |
| 4 | 27/08/19 | 28/11/14 |
| 5 | 03/03/19 | 28/11/14 |
| 6 | 01/03/19 | 08/05/10 |
| 7 | 14/07/19 | 15/05/10 |
| 8 | 07/04/19 | 01/07/05 |
| 9 | 31/10/18 | 19/01/10 |
| 10 | 12/05/19 | 19/04/16 |
| 11* | 2019 | 19/04/16 |
| 11* (H154) | 03/09/19 | 06/04/15 |
| 12 | 04/06/19 | 26/09/15 |
| 13 | 10/08/19 | 24/10/08 |
| 14 | 08/10/19 | 01/07/05 |
| 15 | 12/06/04 | 26/08/96 |
| 16 | 21/08/19 | 31/03/92 |
| 17 | 14/03/19 | 26/09/15 |

*Cluster 11 is paraphyletic with H154 predicted to sit distinctly from the rest of the isolates in the cluster

**Table S5: Details from the first appearance of each CRAB cluster**

| **Cluster** | **Introduction** | **Predicted date of MRCA for all isolates within cluster** | **Nosocomial spread** | **Distinct introductions** | **Cluster** | **Earliest collection from first samples after a patietnt's admission** | **Bed unit or room of first appearance** | | |
| --- | --- | --- | --- | --- | --- | --- | --- | --- | --- |
|  |  |  |  |  |  |  | **Patient and environmental isolates** | **Patient isolates only** | **Environmental isolates only** |
| 1 | Prior to study | 09/07/19 | Yes | Yes | 1 | NA | 6, 22, 24, 25 | 23 | 7, 9, 11, 12, 14, 15, 19, 26 |
| 6 | Prior to study | 01/03/19 | Yes | No | 6 | NA | 28 | ·· | 5, 19, 20, 28 |
| 8 | Prior to study | 07/04/19 | Yes | Yes | 8 | NA | 1 | ·· | ·· |
| 15 | Prior to study | 12/06/04 | Yes | Yes | 15 | NA | ·· | ·· | 18 |
| ST138 | Prior to study | ·· | Sporadic | No | ST138 | NA | ·· | 13 | ·· |
| 3 | Single clonal introduction | 20/08/19 | Sporadic | No | 3 | Yes | ·· | 19 | ·· |
| 4 | Single clonal introduction | 27/08/19 | Yes | No | 4 | Yes | ·· | ·· | 16 |
| 14 | Single clonal introduction | 08/10/19 | Sporadic | No | 14 | No | ·· | 24 | ·· |
| 16 | Diversification prior to introduction | 21/08/19 | Yes | No | 16 | Yes | ·· | 13 | ·· |
| 13 | Diversification prior to introduction | 10/08/19 | Yes | No | 13 | Yes | ·· | 17 | ·· |
| 7 | Diversification prior to introduction | 14/07/19 | Yes | No | 7 | No | ·· | ·· | 13 |
| 12 | Diversification prior to introduction | 04/06/19 | Sporadic | No | 12 | No | ·· | ·· | 25 |
| 10 | Diversification prior to introduction | 12/05/19 | Yes | No | 10 | Yes | ·· | ·· | 7 |
| 17 | Diversification prior to introduction | 14/03/19 | Yes | Yes | 17 | Yes | ·· | ·· | 6, 7, 8 |
| 5 | Diversification prior to introduction | 03/03/19 | Yes | No | 5 | Yes | ·· | ·· | 13, 14 |
| 9 | Diversification prior to introduction | 31/10/18 | Yes | No | 9 | Yes | ·· | 18 | ·· |
| 2 | Diversification prior to introduction | 28/05/17 | Yes | Yes | 2 | Yes | ·· | 16 | ·· |
| 11 | Unclear | 2019 | Yes | Yes | 11* | No | ·· | ·· | 1 |
| ST46 | Unclear | ·· | Sporadic | No | ST46 | No | ·· | 23 | ·· |
| ST1554 | Unclear | ·· | Sporadic | No | ST1554 | No | ·· | ·· | Cleaning room |

**Table S6: Distribution of CRAB clusters within ICU locations and patients**

Count of isolates and distinct bed units, rooms and associated-patients (patient or the bed unit that they occupy) that isolates from each cluster was found in. Note, in the intensive care unit there are total of 28 bed units amongst 13 rooms, and a further 4 rooms comprising the sampled common areas; 140 patients or their bed unit were sampled during the study

| **Cluster** | **# isolates** | **# distinct bed units** | **# distinct rooms** | **# distinct patients associated** |
| --- | --- | --- | --- | --- |
| **1** | 224 | 27 | 14 | 49 |
| **2** | 46 | 10 | 5 | 12 |
| **3** | 1 | 1 | 1 | 1 |
| **4** | 6 | 3 | 3 | 3 |
| **5** | 8 | 4 | 2 | 3 |
| **6** | 12 | 2 | 2 | 2 |
| **7** | 3 | 2 | 1 | 2 |
| **8** | 86 | 19 | 10 | 21 |
| **9** | 56 | 13 | 7 | 21 |
| **10** | 15 | 7 | 5 | 8 |
| **11** | 13 | 5 | 4 | 5 |
| **12** | 3 | 1 | 1 | 1 |
| **13** | 31 | 10 | 7 | 11 |
| **14** | 2 | 2 | 2 | 1 |
| **15** | 4 | 3 | 2 | 4 |
| **16** | 19 | 5 | 4 | 4 |
| **17** | 18 | 6 | 3 | 6 |
| **ST138** | 1 | 1 | 1 | 1 |
| **ST1554** | 1 | 1 | 1 | 1 |
| **ST46** | 2 | 2 | 2 | 1 |

**Table S7: Dissemination of CRAB clusters between adjacent bed units and rooms**

Frequency of routine sampling occasions for all rooms containing bed units when: the rooms were sampled, carbapenem-resistant *Acinetobacter baumannii* (CRAB) was present in the room, isolates from the same CRAB cluster was present in multiple bed units (BU) of the room and isolates from the same CRAB cluster were present in one or more adjacent rooms. Separate evaluations were undertaken for all isolates collected, environmental isolates only (i.e. potential direct environment-environment transmission). and patient isolates only (i.e. potential direct patient- patient transmission). Percentages are calculated as a proportion of occasions when a room was sampled.

|  | **Occasions when a room was sampled** | **Occasions when a room contained CRAB** | **Occasions when multiple BU within a room contained the same CRAB cluster** | **Occasions when a room was adjacent to another with the same CRAB cluster** |
| --- | --- | --- | --- | --- |
| **All isolates** | 156 | 117 (75·00 %) | 46 (38·66 %; 11 clusters) | 76 (48·72 %; 8 clusters) |
| **Environmental isolates** | 156 | 112 (71·79 %) | 45 (34·35 %; 10 clusters) | 69 (44·23 %; 8 clusters) |
| **Patient isolates** | 154 | 63 (40·91 %) | 3 (2·73 %; 2 clusters) | 8 (5·19 %; 2 clusters) |

**Table S8: Potential transfer of CRAB associated with consecutive patients occupying a bed unit**

The presence of the same CRAB cluster associated with consecutive patients who occupy a bed unit is indicative that cleaning of the bed unit between admissions has been unsuccessful. Occasions when one patient left a bed unit and the next was admitted or the bed unit was left unoccupied were defined as “consecutively sampled patients”. Potential transfer events were counted when the same CRAB cluster was present in the bed unit before and after the arrival of the new patient in “donor” and “recipient” weeks, respectively. The numbers of consecutively sampled patients and transfer events were determined considering a number of circumstances: 1) when any samples were taken in each week of sampling, 2) when environmental and patient screening samples were taken associated from both the “donor” and “recipient” weeks of sampling, 3) when environmental samples were taken from both the “donor” and “recipient” weeks of sampling (i.e. transmission may be from environmental to environmental sites) and 4) when patient screening samples were taken from both the “donor” and “recipient” weeks of sampling (i.e. transmission may be directly from patient to patient). On two occasions, two patients were recorded in same bed unit on the same date as the environment was sampled before, the patient changed and then the patients were screened later in the day. Each patient was counted as part of consecutive patient pairs with incomplete sampling. None of the samples taken from these bed units on these occasions yielded CRAB. Some patients were admitted to multiple bed units during the course of the study; when these patients were recipient to a CRAB transfer event, none of these patients had the same CRAB clusters in their former bed unit elsewhere in the ward as in the recipient week (i.e. the CRAB cluster had not been carried with them as they moved beds).

|  | **# consecutively sampled patients** | **Total # potential transfer events detected** | **% patient changes that result in the same CRAB cluster associated with consecutive patients in a bed unit** |
| --- | --- | --- | --- |
| Any samples taken (1) | 150 | 23* | 15·33 |
| Complete sample sets (2) | 116 | 16 | 13·79 |
| Environmental sampling (3) | 146 | 23 | 15·75 |
| Patient screening (4) | 94 | 0 | 0·00 |

*For every potential transfer event, the transferred CRAB cluster was found in the environmental samples of the bed unit on both the donor and recipient sampling dates. On five occasions (see table S11), the transferred CRAB cluster was also found in patient screening sample on the former sampling date and not the latter sampling date, though on two of these occasions, patient screening samples were not taken on the latter date. On one occasion, the transferred CRAB cluster was also found in the patient screening samples on the latter date, suggesting that the patient acquired CRAB from insufficient cleaning of the bed unit between patients.

**Table S9: Details of potential CRAB transfer events associated with consecutive patients in the same bed unit**

For each potential transfer event, the bed unit, donor week and CRAB cluster involved are specified. Details describe whether the same cluster was found in the same room or adjacent rooms in the donor week which would suggest that the CRAB acquisition may have been from other donors rather than due to consecutive patients in the bed. Additional information indicates if CRAB was identified in patient samples in the donor or recipient week indicating whether this influences transmission and if patients are acquiring CRAB from unsuccessful terminal cleaning.

| **Bed unit #** | **Donor week #** | **CRAB cluster** | **Same cluster found in same room** | **Same cluster found in adjacent room** | **Includes CRAB from patient screening in donor week** | **Includes CRAB from patient screening in recipient week** |
| --- | --- | --- | --- | --- | --- | --- |
| 7 | 7 | 1 | Yes | Yes | ·· | ·· |
| 7 | 11 | 10 | ·· | ·· | ·· | ·· |
| 7 | 12 | 10 | ·· | Yes^*^ | ·· | ·· |
| 8 | 10 | 1 | Yes | ·· | ·· | ·· |
| 12 | 10 | 2 | Yes | ·· | ·· | ·· |
| 12 | 11 | 2 | Yes | ·· | ·· | ·· |
| 12 | 12 | 2 | Yes | ·· | ·· | ·· |
| 14 | 10 | 1 | ·· | ·· | ·· | ·· |
| 15 | 7 | 8 | Yes | Yes | ·· | ·· |
| 16 | 10 | 2 | Yes | ·· | ·· | ·· |
| 17 | 2 | 9 | ·· | Yes | ·· | ·· |
| 18 | 3 | 9 | Yes | Yes | Yes | ·· |
| 19 | 2 | 9 | Yes | Yes | ·· | ·· |
| 19 | 3 | 9 | Yes | Yes | ·· | ·· |
| 20 | 11 | 1 | ·· | Yes | ·· | ·· |
| 20 | 12 | 1 | Yes | Yes | ·· | Yes |
| 21 | 12 | 1 | Yes | Yes | ·· | ·· |
| 22 | 2 | 1 | Yes | Yes | Yes | ·· |
| 23 | 4 | 1 | ·· | Yes | Yes^+^ | ·· |
| 23 | 12 | 9 | Yes | Yes | Yes | ·· |
| 24 | 2 | 1 | Yes | Yes | Yes^+^ | ·· |
| 24 | 4 | 13 | ·· | Yes | ·· | ·· |
| 25 | 12 | 1 | Yes | ·· | ·· | ·· |

^*^only associated with patient that moves beds

^+^ patient was not sampled in the recipient week

**Table S10: Plasmids in the DETECTIVE CRAB collection**

| **Plasmid** | **Type** | **Size (bp)** | **Antibiotic resistance genes** | **Reference from** | **GenBank accession** |
| --- | --- | --- | --- | --- | --- |
| pDETAB1 | GR24 | 103,751 | ·· | DETAB-P2 | CP047974 |
| pDETAB2 | GR34 | 100,072 | *bla*_NDM-1_, *ble*_MBL_, *bla*_OXA-58_, *aacC2d*, *sul2*, *msr*(E)-*mph*(E)*, tet*(39) | DETAB-P2 | CP047975 |
| pDETAB3 | Aci2b | 9,132 | ·· | DETAB-P2 | CP047976 |
| pDETAB4 | GR24 | 113,682 | *sul2*, *tet*(B) | DETAB-E227 | CP072527 |
| pDETAB5 | GR13 | 97,035 | *bla*_NDM-1_, *ble*_MBL_, *bla*_OXA-58_, *aacC2d*, *sul2*, *msr*(E)-*mph*(E) | DETAB-E227 | CP072528 |
| pDETAB6 | Aci3 | 7,145 | ·· | DETAB-E227 | CP072529 |
| pDETAB7a  pDETAB7b  pDETAB7c^1^  pDETAB7d | Aci6 | 72,258  73,444  84,261  84,262 | ··  ··  ··  ·· | DETAB-E158  DETAB-P65  ··  DETAB-P43 | CP077838  CP077836  ··  CP077833 |
| pDETAB8 | Aci6 | 70,426 | ·· | DETAB-P24 | CP077847 |
| pDETAB9a  pDETAB9b | Aci9 | 11,194  11,193 | ··  ·· | DETAB-P90  DETAB-E50 | CP077842  CP077831 |
| pDETAB10 | Aci1 | 8,731 | ·· | DETAB-P24 | CP077834 |
| pDETAB11 | Aci2a | 5,602 | ·· | DETAB-E350 | CP077829 |
| pDETAB12 | GR24 | 110,967 | ·· | DETAB-E107 | CP077827 |
| pDETAB13 | GR13 | 91,083 | ·· | DETAB-P39 | CP073061 |
| pDETAB14 | novel^2^ | 38,952 | ·· | DETAB-E154 | CP077844 |
| pDETAB15 | GR24 | 112,130 | ·· | DETAB-P16 | CP077849 |
| pDETAB16 | Aci6 | 70,789 | ·· | DETAB-P90 | CP077841 |

^1^ Existence of pDETAB7c is inferred from BLAST query analysis that revealed plasmids that contain the same insertion as pDETAB7d but lack pDETAB7d’s backbone nucleotide polymorphisms. The size listed for pDETAB7c is predicted based on the size of pDETAB7d. As a complete sequence of pDETAB7c was not obtained, a reference isolate and GenBank accession are not listed.

^2^ pDETAB14 does not contain a recognisable *rep* gene and therefore cannot be assigned to a Rep group according to established typing protocols for *Acinetobacter* plasmids. No plasmids closely related to pDETAB14 are found in the GenBank nucleotide database (last search August 3, 2021). The most closely related sequences in GenBank belong to other *Acinetobacter* plasmids, which match a total of approximately 14,000 bp of pDETAB14 with an overall nucleotide identity of approximately 77%.

**Table S11: Signature sequences used for detection of mobile genetic elements and particular configurations of elements in the DETECTIVE CRAB collection**

| **Target** | **Signature sequences^1^** |
| --- | --- |
| Tn*2006* in AbGRI1 | Tn2006_AbGRI1-left; Tn2006_AbGRI1-right |
| Tn*2009* chromosomal position 1 | Tn2009_ST23-left; Tn2009_ST23-right |
| Tn*2009* chromosomal position 2 | Tn2009_H138-left; Tn2009_H138-right |
| Tn*2009* chromosomal position 3 | Tn2009_B99-left; Tn2009_B99-right |
| Tn*2009* chromosomal position 4 | Tn2009_H107-left; Tn2009_H107-right |
| pDETAB7a | Tn6022_uninterrupted; pDETAB7a_bbSNP |
| pDETAB7b | Tn6022_uninterrupted; pDETAB7a_bbSNP; Aci6_ISAba1 |
| pDETAB7c | Tn6022_left; Tn6022_right; pDETAB7a_bbSNP |
| pDETAB7d | Tn6022-left; Tn6022-right; pDETAB7d_bbSNP |
| pDETAB8 | Tn6002_uninterrupted; pDETAB8_region |
| pDETAB16 | Tn6022_uninterrupted; pDETAB16_region |

^1^ “Signature sequences” that span insertion junctions, uninterrupted insertion target sites or characteristic plasmid backbone regions were derived from hybrid-assembled complete genomes. Signature sequences were used to query a database comprised of all contigs from all 551 DETECTIVE CRAB draft genomes. A target was determined to be present in a given genome if that genome yielded complete matches to the characteristic signature sequences for that target. All signature sequences are listed in the supplementary file called “signature-sequences.fa”.
