## Supplementary figures for "Extensive acquisition of carbapenem-resistant *Acinetobacter baumannii* in Intensive Care Unit patients is driven by widespread environmental contamination"

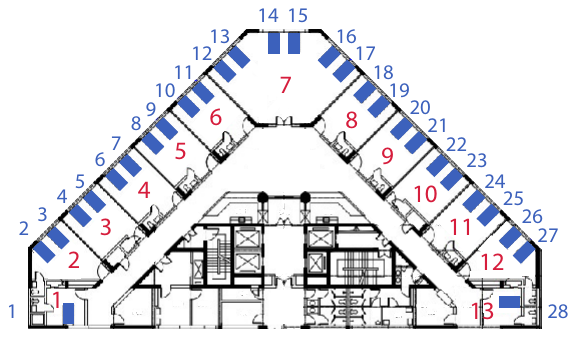

**Figure S1: Map of the ICU**

Architect’s drawing of the intensive care unit at Sir Run Run Shaw Hospital, Hangzhou showing the 13 bedrooms in the ward (numbered red) and the 28 bed units (blue). The communal spaces are in the centre of the ward.

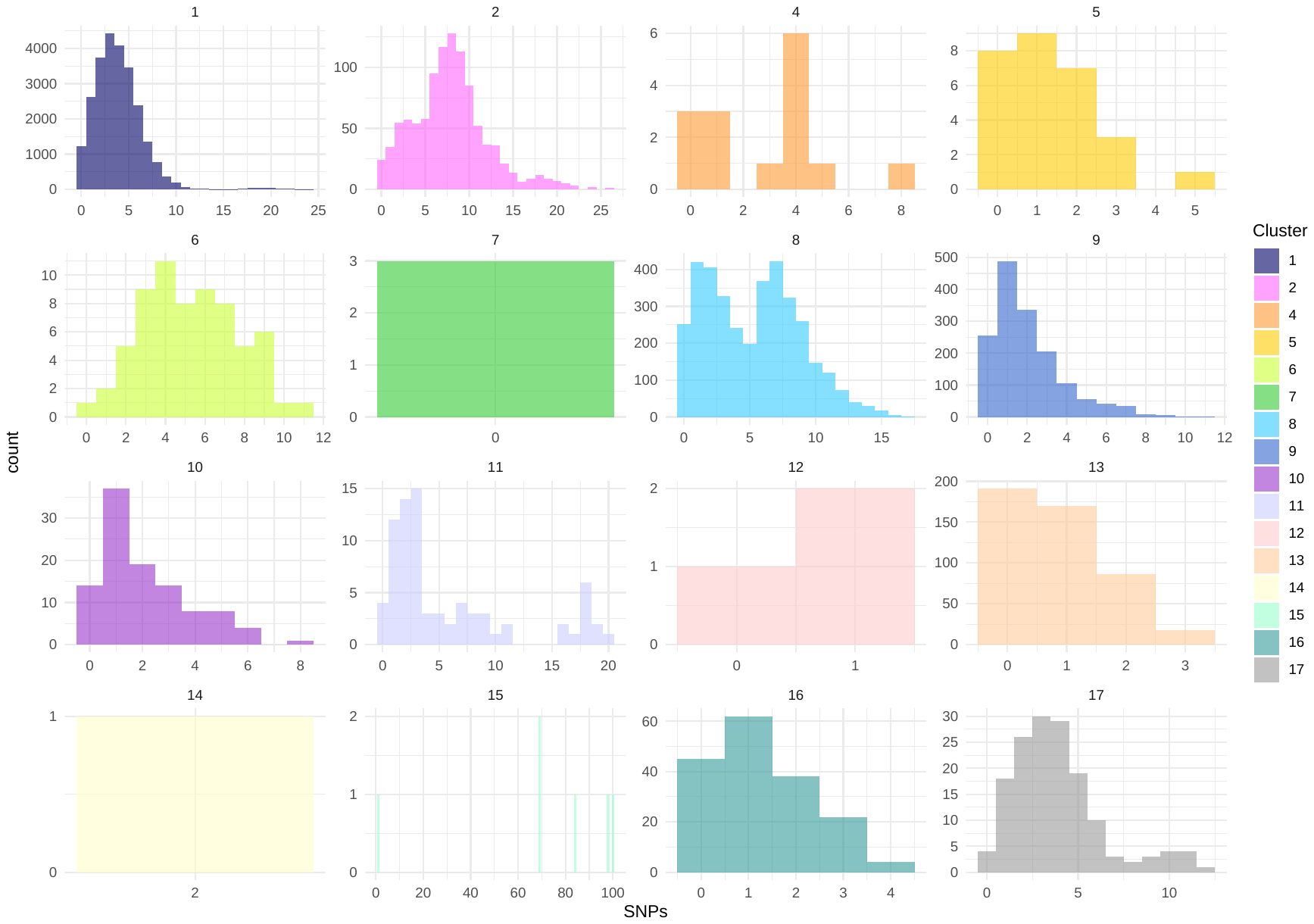

**Figure S2: Core-genome SNP distance histograms for GC2/ST187 clusters with more than one isolate.** SNP distances were calculated across the whole GC2/ST187 population and the frequency distribution of the SNP-distances within each cluster was visualised in each panel, with the panels coloured according to corresponding cluster. SNP distances can be found on x-axes; noting that the axis scale varies between panels. The frequency of each SNP distance within a cluster is on the y-axes, also noting varying scales.

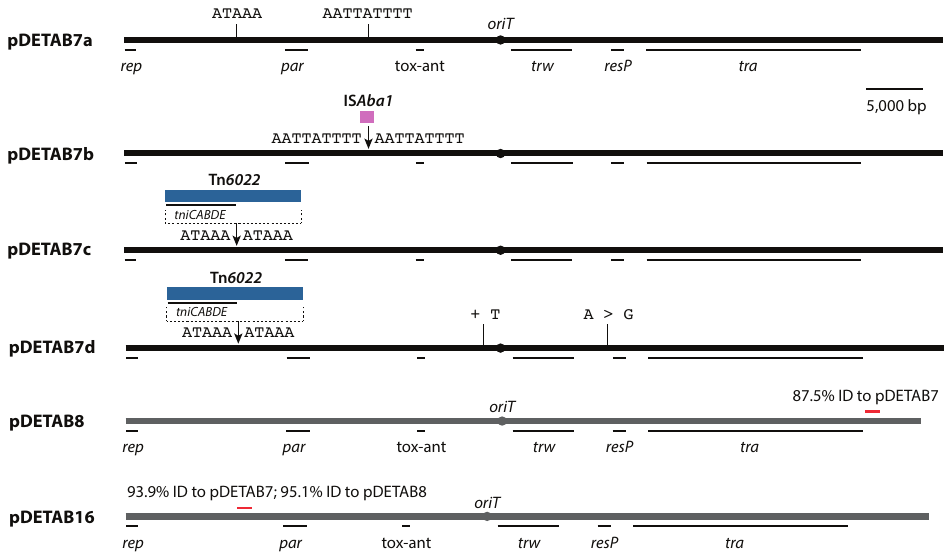

**Figure S3: Aci6 plasmids.** Linear, scaled maps of Aci6 plasmids found in this collection. Plasmid backbones are shown as vertical lines with the locations of named, putative replication, maintenance and transfer determinants indicated below. The putative origin-of-transfer (*oriT*) is shown as a labelled circle. Insertions in plasmid backbones are indicated above, with the sequences of insertion sites and target site duplications shown. The positions and identities of single nucleotide polymorphisms are also indicated above. The regions used to distinguish pDETAB8 and pDETAB16 backbones from one another and from pDETAB7 are indicated by red lines. The nucleotide identities of the regions indicated by red lines relative to the other backbone types are are labelled. The sequence marked by a red line in pDETAB8 is identical to the corresponding sequence in pDETAB16.
